# Activation of the apolipoprotein A1-SRB1 pathway increases vascular permeability in ovaries with lipotoxicity

**DOI:** 10.1101/2025.11.04.25339539

**Authors:** Hyeri Park, Jin Seok, Dae Hyun Lee, Hee Won Jang, Jae Young Shin, Jong-Uk Namkung, Hankyu Lee, Jung Ryeol Lee, Gi Jin Kim

## Abstract

**Objective:** Excessive cholesterol causes lipid accumulation and plaque buildup in blood vessels, reducing blood supply and ovarian function. The apolipoprotein A1 (ApoA1)-scavenger receptor class B member 1 (SRB1) signaling is crucial in lipid-induced vascular permeability and ovarian steroidogenesis. We previously reported that placenta-derived mesenchymal stem cells (PD-MSCs) improve ovarian function through glucose metabolism and antioxidant effects in a metabolic disorder rat model treated-TAA. However, their therapeutic correlation between ovarian and vascular functions remains unclear. In this study, we analyzed whether PD-MSCs activate ApoA1-SRB1 signaling in a metabolic disorder rat model, and whether their activation modulated vascular permeability and thereby alleviatees ovarian dysfunction.

**Approach & Results:** The ovarian metabolic disorder model was established by intraperitoneal injection of thioacetamide (TAA), a lipid toxicity inducer, twice weekly for 12 weeks. After 8 weeks of TAA administration, PD-MSCs (2x10^6^ cells) were transplanted via tail vein. After sacrificed at 12 weeks, the expressions of factors for vascular permeability and follicular development were analyzed. PD-MSC transplantation (Tx) significantly decreased lipid accumulation in both blood and ovarian tissues. Additionally, increased ApoA1 expression induced by PD- MSCs triggered SRB1 activation and decreased vascular permeability via eNOS expression in ovarian tissues from patients with metabolic diseases (\**p*<0.05). Furthermore, the expression of genes related to vascular permeability (e.g., eNOS and Erg-3) and steroidogenesis was significantly greater in the PD-MSC Tx group than in the sham group (\**p*<0.05).

**Conclusions:** These findings suggest that PD-MSCs normalize vascular function via the ApoA1-SRB1 pathway in ovarian diseases associated with lipid toxicity, suggesting a novel strategy for treating reproductive disorders linked to metabolic dysfunction.

**Highlights:**

- PD-MSCs significantly decreased lipid accumulation in both blood and ovarian tissues, alleviating lipid-induced ovarian dysfunction.
- PD-MSCs enhanced ApoA1-SRB1 signaling, which activated eNOS expression and reduced vascular permeability in ovarian tissues.
- PD-MSC treatment improved vascular and steroidogenic gene expression, improving ovarian function in metabolic disorder models.

## INTRODUCTION

Metabolic diseases are increasing across a wide range of age groups due to various environmental factors, such as westernized dietary habits, stress, lack of sleep, and sedentary lifestyles ^1^. The liver, a major site in metabolism, plays a central role in all metabolic processes by breaking down fats and generating energy ^2^. Moreover, metabolic dysfunction in the liver is directly linked to reproductive health, as metabolic imbalance affects ovarian function through systemic metabolic regulation. The liver serves as the metabolic hub of the body and is involved in the detoxification of steroid hormones, including cholesterol transport and estrogen synthesis. Therefore, liver pathology may contribute to female endocrine disorders *via* the liver‒ovary axis ^3^.

Polycystic ovary syndrome (PCOS), a common metabolic and endocrine disorder in premenopausal women, is one of the most prevalent metabolic and endocrine abnormalities among premenopausal women. Several symptoms of PCOS resemble those observed in perimenopausal individuals ^4^. The onset criteria for PCOS are multifactorial in nature, but the diagnostic criteria are typically categorized into four main aspects: (1) metabolic dysfunction, (2) excess androgen levels, (3) polycystic ovarian morphology, and (4) menstrual irregularities ^5^. Metabolic syndrome in PCOS patients is diagnosed when at least three of the following five risk factors are present: abdominal obesity (>88 cm), high triglycerides (≥150 mg/dl), low high-density lipoprotein cholesterol (HDL-C) (<50 mg/dl), high blood pressure (≥130/85 mmHg), and impaired fasting glucose (110–126 mg/dl) 11368702 ^6^. Wild et al. reported that women with PCOS present elevated low-density lipoprotein cholesterol (LDL-C) levels independent of BMI, along with abnormal HDL-C function rather than simply reduced HDL-C levels ^7^. Thus, the functional quality of HDL-C, rather than just its concentration, is now considered a crucial therapeutic target in metabolic disorders, such as PCOS ^8^.

Lipid regulation in the ovary influences steroidogenesis by controlling cholesterol availability, which is primarily mediated by steroidogenic acute regulatory protein (StAR) and sex hormone-binding globulin (SHBG), which affect steroid hormone transport and bioavailability ^9^. Excessive lipid accumulation disrupts this process, leading to impaired hormone production and ovarian dysfunction. Conversely, proper lipid metabolism supports balanced steroidogenesis, which is essential for normal reproductive function ^10^.

The characteristics of metabolic disorders lead to abnormal vascular function ^11^. Excessive cholesterol, an important component of the body’s lipid profile, leads to the accumulation of lipids, triggering metabolic disorders. This accumulation of lipids leads to the buildup of plaque within the blood vessels, resulting in inadequate blood supply from the arteries and subsequent organ damage ^12^. Vascular dysfunction not only affects systemic circulation but also plays a critical role in ovarian function, as ovarian vascularization is essential for follicular growth and hormonal regulation. Women with PCOS experience a decrease in ovulation rates due to abnormalities in ovarian angiogenesis ^13^. The ovarian vasculature is a crucial factor in ovarian function, as it delivers nutrients, cytokines, and hormones to follicles, thereby regulating follicular development and maturation ^14^. The ovarian vasculature infiltrates theca cells but does not penetrate the basement membrane or granulosa cells. Instead, granulosa cells and oocytes receive nutrients, hormones, and cytokines through diffusion across the theca cell layer ^15^. Damage to the ovarian vascular network, a critical factor in regulating ovarian function, results in nutrient deprivation of the follicles ^16^. Battaglia C was the first to report that ovarian blood flow in patients with PCOS is increased compared with that in controls ^17^. Pietro and colleagues reported that women with PCOS exhibit abnormal ovarian blood flow and disruption of vascular angiogenesis ^13^. However, previous studies have focused primarily on angiogenesis without considering vascular permeability and endothelial homeostasis, which are equally important for ovarian function. Tahergorabi and colleagues proposed studying ovarian angiogenesis imbalance as a new tool for diagnosing and managing PCOS on the basis of evidence that VEGF-dependent angiogenesis is closely associated with BMI, further linking metabolic imbalance to ovarian vascular dysfunction ^18^. The association between metabolic disorders and endothelial cell dysfunction in individuals with PCOS is well known. Oncul and colleagues demonstrated that women with PCOS experience endothelial dysfunction due to hyperandrogenism, regardless of BMI, as evidenced by the correlation with serum soluble lectin-like oxidized low-density lipoprotein receptor-1 (sLOX-1) ^19^.

Although various pharmacological therapies are available for treating PCOS, targeting the broad spectrum of PCOS symptoms causes challenges, and each medication has its own set of side effects, such as menstrual bleeding and cramping ^20^. To overcome the limitations of pharmacotherapy, many scientists are researching mesenchymal stem cell (MSC) therapies for PCOS. According to Kalhori et al., the intravenous transplantation of bone marrow-derived MSCs (BM-MSCs) normalizes follicular development through their anti-inflammatory, antioxidant and antiapoptotic effects in a testosterone enanthate-induced PCOS mouse model ^21^. Xie and colleagues reported that intraovarian transplantation of umbilical cord-derived MSCs (UC-MSCs) alleviates ovarian dysfunction by inhibiting the inflammatory response in a dehydroepiandrosterone (DHEA)-induced PCOS mouse model ^22^.

Furthermore, MSC-based therapies for PCOS are being investigated in *in vitro* experiments. Chung and colleagues reported that BM-MSCs regulate steroidogenesis and androgen production in primary theca cells ^23^. According to Zhao et al., UC-MSC-derived exosomes reduce granulosa cell inflammation by inhibiting NF-kB signaling in PCOS patients ^24^. In previous reports, we demonstrated that placenta-derived MSC (PD-MSC) transplantation restores follicular development through vascular remodeling by VEGF and PDGF signaling in an ovariectomized rat model ^25, 26^. Furthermore, PD-MSC transplantation normalized ovarian function by regulating glucose metabolism in a thioacetamide (TAA)-induced lipid toxicity model, further linking lipid metabolism to ovarian dysfunction ^27^. However, despite growing evidence supporting the benefits of MSC therapy, the exact mechanisms underlying their regulation of ovarian vascular permeability remain unclear.

Apolipoprotein-A1 (Apo-A1), the primary protein component of discoidal HDL, binds to HDL as a lipid component and facilitates the import of cholesterol while regulating enzymatic modification to transport mature HDL ^28^. Interestingly, ApoA1 not only is a key regulator of lipid metabolism but also exerts anti-inflammatory and vasoprotective effects, making it a promising therapeutic target for metabolic disorders that induce PCOS. Additionally, scavenger receptor-B1 (SR-B1), a known receptor or HDL, is closely associated with female fertility. Miettinen et al. reported that mice deficient in SR-B1 exhibit abnormal lipoprotein metabolism and reversible female infertility ^29^.

Given the dual role of ApoA1-SRB1 signaling in vascular permeability and ovarian function, we hypothesize that PD-MSC transplantation can activate this pathway to normalize ovarian function in metabolic disorder models. Therefore, the aim of this study was to determine whether PD-MSC-mediated activation of the ApoA1-SRB1 pathway can regulate vascular function and steroidogenesis, ultimately alleviating ovarian dysfunction in PCOS models.

## MATERIALS AND METHODS

### Cell culture

The use of human placental samples for research purposes was approved by the Institutional Review Board of CHA Gangnam Medical Center, Seoul, Republic of Korea (IRB-07-18). In brief, as outlined in our previous report, human PD-MSCs were isolated from normal placental chorionic plates at term (gestation ≥ 37 weeks) without any medical, obstetrical or surgical complications ^30^. PD-MSCs were cultured in alpha-minimum essential medium (α-MEM; HyClone, Utah, USA) supplemented with 10% fetal bovine serum (FBS; Gibco-BRL, Oklahoma, USA), 1% penicillin/streptomycin (P/S; Gibco-BRL), 1 μg/ml heparin (Sigma‒Aldrich, Missouri, USA) and 25 ng/ml FGF-4 (PeproTech, New Jersey, USA) in a 37°C incubator in a humidified atmosphere of 5% CO_2_. Harvested PD-MSCs were labeled with a PKH67 Fluorescent Cell Linker Kit (Sigma‒Aldrich). Then, 2×10^6^ PD-MSCs were injected intravenously into the transplanted group.

### Construction of the TAA-induced rat model

The Institutional Animal Care and Use Committee (IACUC-220044) of CHA University, located in Seongnam-si, Republic of Korea, approved the animal models and experimental protocols. Seven-week-old female Sprague‒Dawley rats (Koatech, Pyeongtaek-si, Republic of Korea) were housed in an air-conditioned animal facility. The rats were housed in pairs in plastic cages containing corn cob bedding and were provided with unlimited access to standard commercial food and tap water. The rats were maintained under standard conditions with a temperature of 21°C and a 12-hour light‒dark cycle. The ovarian metabolic dysfunction model was constructed via the intraperitoneal injection of 150 mg/kg TAA (Sigma‒Aldrich), which induces lipid toxicity, twice a week for 12 weeks. Eight weeks after intraperitoneal injection, 2×10^6^ PD-MSCs labeled with the PKH67 Fluorescent Cell Linker Kit were transplanted intravenously through the tail vein. All the rats were euthanized 4 weeks after transplantation, and blood and ovarian tissue samples were collected.

### 70-kDa lysinated rhodamine dextran

Analysis of 70-kDa lysinated rhodamine dextran (Thermo Fisher Scientific) was performed to evaluate the vascular permeability *in vivo* as described previously, with slight modifications ^31^. The last day of TAA-induced rat model construction, 1.2 mg of 70-kDa lysinated rhodamine dextran in 100 µl distilled water (D.W.) was intravenously injected into the tail vein of each group of rats. At 10 min post-injection, all groups of rats were sacrificed. The ovarian tissues were collected, and the frozen ovarian section blocks were fixed with 4% paraformaldehyde for 10 min. Then, blood vessels were visualized by staining with an anti- CD31 antibody as described for immunofluorescence. The tissues were counterstained and mounted with mounting medium containing DAPI (Vectashield, Burlingame, USA). A fluorescence microscope (Zeiss LSM 680, Oberkochen, Germany) was used to observe the tissues at 63x magnification. Every section of each slide was observed, and representative images were captured.

### Isolation of primary theca cells

Seven-week-old female Sprague‒Dawley rats were sacrificed using a carbon dioxide atmosphere. Rat ovarian tissues were collected and rinsed with saline and DPBS containing 1% penicillin/streptomycin. After being rinsed, the ovarian tissues were briefly incubated with McCoy’s 5A medium supplemented with 1% penicillin/streptomycin and 2 mM L-glutamine. Two to three ml of medium was dispensed onto a glass Petri dish, and the ovarian tissues were punctured with a 26-gauge needle for 15 min. After puncturing, the ovarian tissues and media from the glass Petri dish were transferred to a 100 μm cell strainer. The remaining ovarian tissues on the cell strainer were transferred to a clean tube, enzyme medium (Hanks’ balanced salt solution containing 10 mg/ml BSA, 50 μg/ml DNase I and 4 mg/ml collagenase) was added, and the samples were incubated in a 37°C incubator in a humidified atmosphere of 5% CO_2_ for 1 h. After incubation, the ovarian tissues and enzyme medium were transferred to a 40 μm cell strainer, and McCoy’s 5A medium supplemented with 5% FBS, 1% penicillin/streptomycin and 2 mM L-glutamine was added. The filtrate obtained through the strainer was centrifuged at 1000 rpm for 5 min, and the supernatant was removed. After cell counting, the separated theca cells were seeded in McCoy’s 5A medium supplemented with 1% penicillin/streptomycin and 2 mM L-glutamine. After 24 h of incubation, when the cells had attached, the culture medium was replaced with McCoy’s 5A medium containing 5% FBS, 1% penicillin/streptomycin, and 2 mM L-glutamine.

### Induction of superovulation

Nineteen-week-old female rats were divided into three groups: a normal group, a TAA-induced group, and a TAA-induced group with PD-MSC transplantation. PMSG (Market Livestock Supplies Co., Ltd., Yongin-si, Republic of Korea) and hCG (Sigma‒ Aldrich) were dissolved in physiological saline. Female rats received 300 IU/kg PMSG via intraperitoneal injection at 20:00, followed by 300 IU/kg hCG via intraperitoneal injection 50 h later. After 24 h, all female rats were euthanized, and their oviduct tissues were collected.

### Low-temperature SDS‒PAGE

SDS‒PAGE was conducted at a low temperature to determine the concentrations of the active (dimer) and inactive (monomer) forms of the eNOS enzyme ^32^. Total protein was incubated in 1X Laemmli buffer without 2.5% β-mercaptoethanol (β-ME) at 37°C for 5 min. The samples were subsequently subjected to SDS‒PAGE via 6–8% gels at 4°C. Before electrophoresis, the gels and buffers were equilibrated at 4°C, and during electrophoresis, the buffer tank was kept in a cold room at 4°C to ensure that the gel temperature remained below 15°C. The separated proteins were transferred to PVDF membranes (Bio-Rad Laboratories, California, USA) at 100 V for 100 min in a cold room at 4°C. Then, the membrane was blocked with 5% BSA for 3 h before being incubated for 20 h at 4°C with a monoclonal anti-eNOS antibody (BD Biosciences, San Jose, USA). Subsequent steps were carried out in a manner similar to that of a standard western blot procedure. A ChemiDoc XRS+ imaging system (Bio-Rad Laboratories) was used to detect the protein bands, and the protein bands were analyzed using the ImageJ program (Wayne Rasband).

### H&E staining for follicle counting

The ovarian tissues were fixed with 10% neutral buffered formalin (BBC, Washington, USA) and embedded in paraffin. The paraffin blocks were serially sectioned into 4-μm ovarian tissue sections. The ovarian tissue sections were deparaffinized by incubation in a 60°C dry oven for 1 h, followed by cooling at room temperature for 1 h. Then, the tissues were deparaffinized with xylene and ethanol. The deparaffinized tissues were rinsed with tap water. The slides were subsequently stained with Harris hematoxylin (Leica Biosystems, Wetzlar, Germany) for 7 min, briefly dipped in 0.1% HCl for 2 secs, and then stained with alcoholic eosin Y solution (Sigma‒Aldrich). The stained slides were scanned for whole ovarian tissues using 3D HISTECH (The Digital Pathology Company, Budapest, Hungary). Follicles were counted by selecting a slide every 100-μm interval from the serially sectioned slides. In accordance with previous studies, follicles were categorized and enumerated as primordial, primary, secondary, antral, or atretic follicles ^33^. The follicle counts were analyzed, and verification of follicle counting was conducted with three or more individuals.

### Nile red staining

Frozen ovarian section blocks were sectioned at a thickness of 7 μm and fixed with 4% paraformaldehyde for 10 min. After air drying, the fixed ovarian tissues were washed with precooled 1X PBS three times for 5 min each at room temperature. A stock solution of 1 mg/ml Nile red (Sigma–Aldrich) was prepared in acetone and subsequently diluted to a concentration of 0.5 μg/ml in 1× PBS. The ovarian tissues were stained with Nile red working solution in a humidified chamber for 1 h. After staining, the ovarian tissues were washed with precooled 1X PBS three times for 5 min each at room temperature. The tissues were counterstained and mounted with mounting medium containing DAPI (Vectashield, Burlingame, USA). A fluorescence microscope (Zeiss Axiocam 506 color, Oberkochen, Germany) was used to observe the tissues at 40x magnification. All parts of each slide were observed, and representative images were captured.

### BODIPY 505/515 staining

Frozen ovarian section blocks were generated, and theca cells and HUVECs were fixed with 4% paraformaldehyde for 10 min. The samples were rinsed three times for 5 min each with precooled 1X PBS. The samples were incubated with a 10 μg/ml BODIPY^®^ 505/515 (Thermo Fisher Scientific) solution for 30 min in a dry oven at 37°C. The samples were rinsed three times for 5 min each with precooled 1X PBS. Then, the samples were counterstained and mounted with mounting medium containing DAPI (Vectashield). A fluorescence microscope (Zeiss Axiocam 506 color, Oberkochen, Germany) was used to observe the tissues at 40x magnification. All parts of each slide were observed, and representative images were captured.

### HUVEC vascular formation

HUVECs were cultured with ECM (Science Cell, California, USA) in a 37°C incubator in a humidified atmosphere of 5% CO_2_. Harvested 3x10^4^ HUVECs were seeded on Matrigel-coated 24-well culture plates. After 24 h, the HUVECs were treated with 70 mM TAA for 24 h. After 24 h, the medium was replaced with ECM without 70 mM TAA. PD- MSCs (1x10^4^) were seeded on 8 μm pore size inserts (Falcon, New York, USA) for 24 h. After cocultivation, the inserts were removed, and the medium was replaced with ECM containing 2 μg/ml Alexa Fluor 488-conjugated Ac-LDL (Invitrogen) for 24 h in a 37°C incubator in a humidified atmosphere of 5% CO_2_. After LDL staining, the HUVECs were washed two times with DPBS. The length of vessel branching was quantified utilizing the ImageJ program.

### HUVEC vascular permeability assay

HUVECs were cultured with ECM (Science Cell, California, USA) in a 37°C incubator in a humidified atmosphere of 5% CO_2_. Matrigel was coated on an insert with a 0.4 μm pore size (Falcon) for 3 h. After coating, 3x10^4^ harvested HUVECs were seeded on a 0.4 μm pore size insert for 24 h. Then, the HUVECs were treated with 70 mM TAA for 24 h. After treatment, the medium was changed to ECM without 70 mM TAA, and PD-MSCs were seeded in 24-well plates (Falcon) for 24 h. At that point, the inserts were transferred to fresh wells containing ECM. ECM medium containing 2 μg/ml dextran (Sigma‒Aldrich) was added to the inserts. After 20 min, the medium from each 24-well plate was transferred to a black 96-well assay plate (Costar, Washington, USA). The plate was analyzed using a Tecan assay to measure the absorbance at 485 nm (excitation) and 535 nm (emission).

### Statistical analysis

All experiments were conducted in duplicate or triplicate. The results are presented as the means ± standard errors. GraphPad Prism 5.0 (GraphPad Software, Inc., CA, USA) was used to conduct statistical analysis using one-way ANOVA followed by Tukey’s multiple comparisons test. *p* values less than 0.05 were considered significant.

## RESULTS

### PD-MSC transplantation reduced lipid accumulation in the ovaries of TAA-injured rats

The weight of the ovary decreases as its function decreases due to conditions such as PCOS and ovarian aging ^34^. To determine whether TAA, a lipid toxicity-inducing agent, causes ovarian dysfunction under conditions such as PCOS and ovarian aging, we measured ovarian weight. TAA was administered at a dose of 150 mg/kg twice a week for 12 weeks, and at week 8, PD-MSCs were transplanted via the tail vein. After the experiment, ovarian tissue samples were collected for analysis (Figure 1A). Compared with that in the normal group, the size of the ovarian tissue in the NTx group was reduced, whereas in the Tx group, it appeared similar in size to that in the normal group (Figure 1B). Compared with that in the normal group, the ovary-to-body weight ratio substantially decreased in the NTx group, whereas it significantly increased in the Tx group (Figure 1C; \**p*<0.05).

**FIGURE 1.**
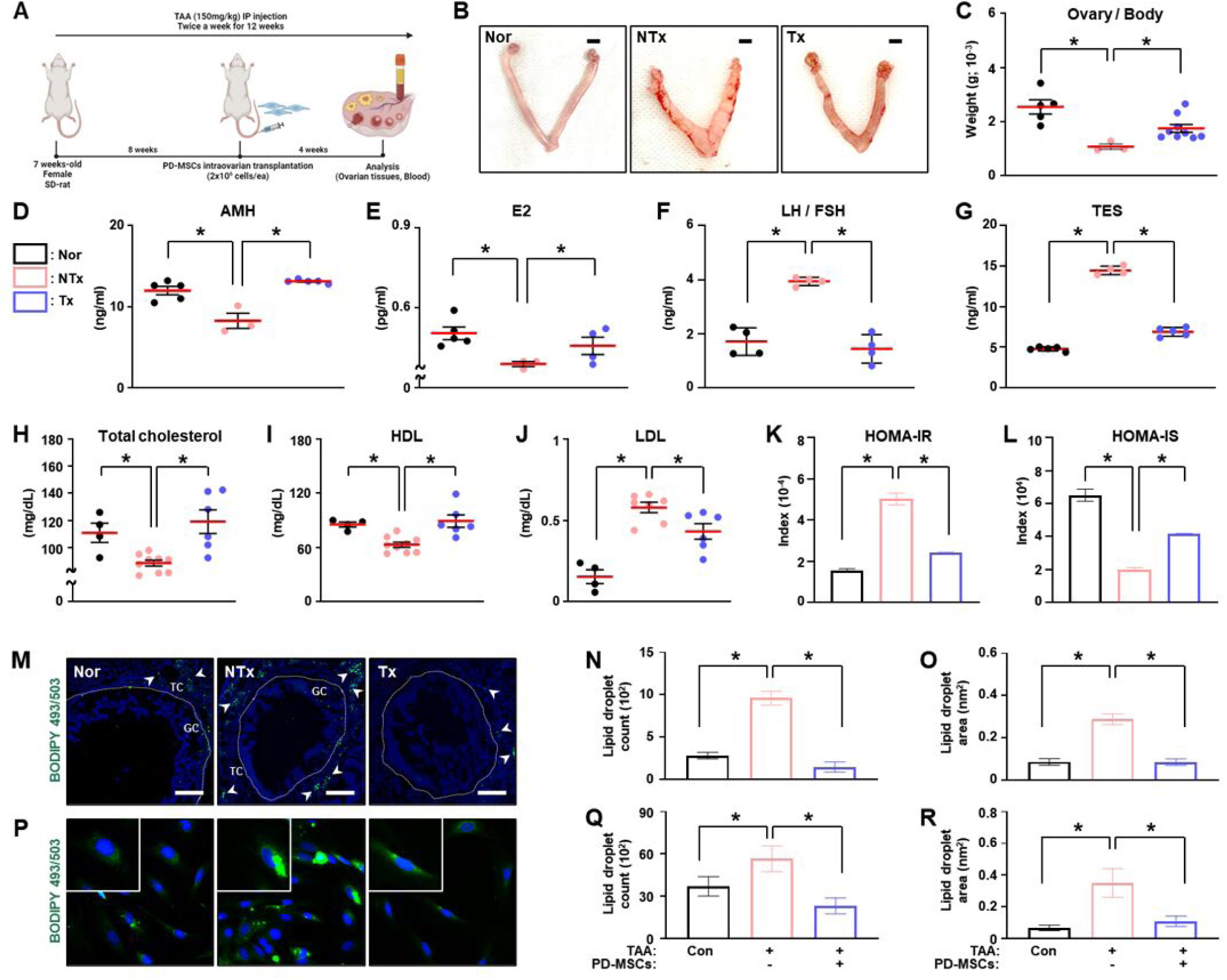
Effect of PD-MSCs on reducing lipid accumulation in ovaries with lipotoxicity. (a) Schematic of the transplantation of PD-MSCs into an ovarian rat model with lipotoxicity. (b) Morphological analysis of the ovarian tissues of TAA-injured rats after sacrifice. (c) The ratio of ovary weight to body weight was measured after the mice were sacrificed. (d) The secretion of AMH, (e) E2, (f) LH, FSH and (g) TES was analyzed by ELISAs. (h) Total cholesterol, (i) HDL-C and (j) LDL-C were analyzed via biochemical analysis. (k) HOMA-IR and (l) HOMA-IS were calculated from insulin and glucose levels. (m) Lipid droplets in ovarian follicles were visualized by BODIPY 505/515 staining. (n-o) The number and area of lipid droplets were quantified via the ImageJ program. Magnification: 40X; scale bar: 50 μm. (p) Lipid droplets in primary theca cells were visualized by BODIPY 505/515 staining. (q-r) The count and area of lipid droplets were quantified via the ImageJ program. Magnification: 40X; scale bar: 50 μm. * *p*<0.05. The *p* value was determined via a nonparametric Kruskal‒ Wallis test followed by Conover‒Iman’s multiple comparisons with BH correlation.

In addition to regulating ovarian function, maintaining the balance of endocrine hormones is essential for follicular development. The level of anti-Mullerian hormone (AMH) in individual serum samples was significantly lower in the NTx group than in the normal group and was significantly greater in the Tx group than in the NTx group (Figure 1D; \**p*<0.05). The level of estradiol (E2) in individual serum samples was significantly lower in the NTx group than in the normal group but significantly greater in the Tx group (Figure 1E; \**p*<0.05). The ratio of luteinizing hormone (LH) to follicle stimulating hormone (FSH) is a well-known marker indicating the absence of ovulation in women with PCOS due to irregular menstrual cycles ^35^. The ratio of LH to FSH in individual serum samples was significantly greater in the NTx group than in the normal group but significantly lower in the Tx group (Figure 1F; \**p*<0.05). Testosterone (TES) is excessively produced in the context of ovarian imbalance caused by PCOS ^36^. The TES levels in individual serum samples strongly increased in the NTx group compared with those in the normal group but significantly decreased in the Tx group, approaching levels similar to those in the normal group (Figure 1G; \**p*<0.05). These results indicate that while metabolic dysfunction impairs ovarian function, the transplantation of PD-MSCs promotes ovarian function.

Cholesterol is a crucial substrate for the synthesis of ovarian sex hormones and plays an important role in follicular development. The levels of total cholesterol and HDL-C were significantly lower in the NTx group than in the normal group but significantly greater in the Tx group (Figure 1H-I; \**p*<0.05). However, compared with that in the normal group, LDL-C was significantly increased in the NTx group but significantly decreased in the Tx group (Figure 1J; \**p*<0.05). To assess insulin resistance and high insulin, we assessed common symptoms of metabolic disorders and glucose and insulin levels in the serum, and the homeostatic model assessment for insulin resistance (HOMA-IR) score was calculated. The HOMA-IR score was significantly greater in the NTx group than in the normal group. This score was significantly lower in the Tx group than in the NTx group (Figure 1K; \**p*<0.05). In contrast, the HOMA-IS score was significantly lower in the NTx group than in the normal group but significantly greater in the Tx group than in the NTx group (Figure 1L; \**p*<0.05).

For analysis of lipid accumulation in ovarian tissue, neutral lipid droplets were stained. Interestingly, lipid droplets were predominantly observed near the theca cell layer of the ovarian follicles (Figure 1M). The number and area of lipid droplets in the theca cell layer of mature follicles were significantly greater in the NTx group than in the normal group but significantly lower in the Tx group than in the NTx group (Figure 1N-O; \**p*<0.05). Since lipid droplets accumulate particularly in the theca cell layer, theca cells were isolated from rat ovaries to analyze lipid accumulation specifically at the cellular level (Figure 1P). The number and area of lipid droplets in theca cells were significantly greater in the TAA-treated group than in the control group but significantly lower in the TAA-treated with PD-MSC cocultivation group than in the TAA-treated group (Figure 1Q-R; \**p*<0.05).

### PD-MSC transplantation regulates vascular structure by ApoA1-SRB1 signaling in the ovaries of TAA-injured rats

Lipid deposits damage endothelial cells and cause structural abnormalities in blood vessels; therefore, we conducted a histological analysis of the vascular structure within the ovarian tissue (Figure 2A). The number of blood vessels did not significantly differ among all the groups; however, artery thickness was significantly greater in the NTx group than in the normal group and returned to levels similar to those of the normal group in the Tx group (Figure 2B-C; \**p*<0.05).

**FIGURE 2.**
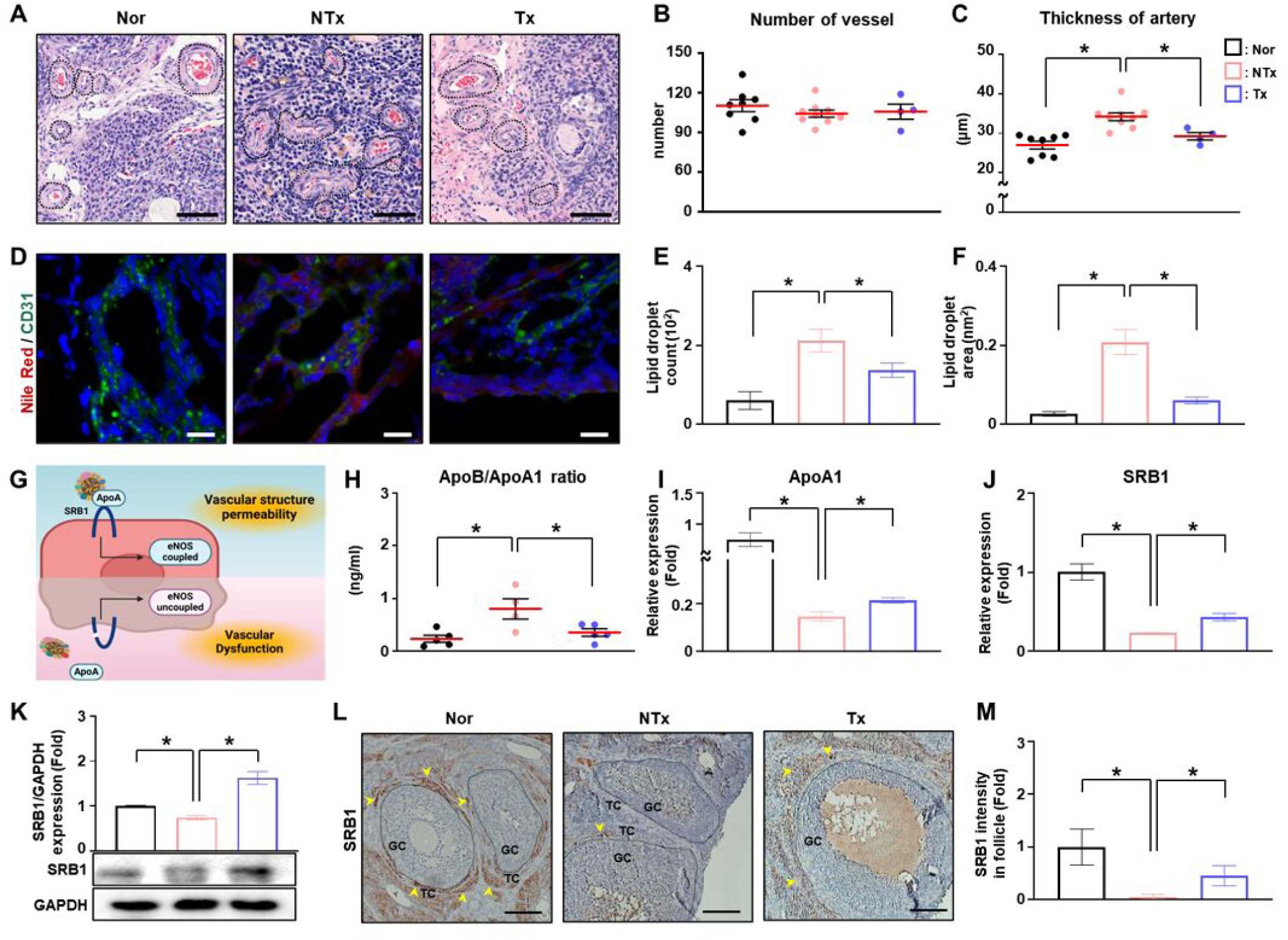
Effect of PD-MSCs on vascular structure by ApoA1-SRB1 signaling in ovaries with lipotoxicity. (a) H&E staining was performed to observe the structure of the ovarian blood vessels. (b) The number of vessels and (c) thickness of the artery were analyzed via the 3D HISTECH program. Magnification: 1.5X, scale bar: 1 mm. (d) Lipid droplets in ovarian blood vessels were visualized by Nile red staining, and endothelial cells were labeled with CD31. (e-f) The count and area of lipid droplets were quantified via the ImageJ program. Magnification: 40X; scale bar: 50 μm. (g) Schematic illustration of the mechanism by which ApoA1-SRB1 signaling promotes eNOS coupling to reduce excessive vascular permeability. (h) The ratio of ApoB to ApoA1 in the serum was analyzed via ELISAs. (i) The mRNA expression of ApoA1 and (j) SRB1 was analyzed by qRT‒PCR. (k) The protein expression of SRB1 was analyzed by western blotting. (l) The localization of SRB1 in ovarian tissues was determined via immunohistochemical staining. (m) The intensity of SRB1 was quantified via the 3D HISTECH program. Magnification: 20X; scale bar: 100 μm. * *p*<0.05. The *p* value was determined via a nonparametric Kruskal‒Wallis test followed by Conover‒Iman’s multiple comparisons with BH correlation.

Lipid accumulation was observed in the theca cell layer of the follicle, where blood vessels permeate and serve as pathways for lipid transport. Therefore, to further investigate lipid accumulation in blood vessels within the ovarian tissue, we performed staining of CD31, a specific marker for blood vessels, along with Nile red staining (Figure 2D). The number and area of lipid droplets in the ovarian blood vessels were significantly greater in the NTx group than in the normal group and significantly lower in the Tx group than in the NTx group (Figure 2E-F; \**p*<0.05). These results suggest that PD-MSCs alleviate lipid accumulation in TAA-induced metabolic disorders.

ApoA-SRB1 signaling is known to regulate vascular structure and permeability (Figure 2G). The ratio of ApoB to ApoA1 is a clinical indicator for metabolic disorders such as PCOS. The ratio of ApoB to ApoA1 in individual serum samples was significantly greater in the NTx group than in the normal group but significantly lower in the Tx group than in the NTx group (Figure 2H; \**p*<0.05). The mRNA expression of ApoA1 was lower in the NTx group than in the normal group but was significantly greater in the Tx group than in the NTx group (Figure 2I; \**p*<0.05). The mRNA and protein expression of SRB1 was significantly lower in the NTx group than in the normal group but significantly greater in the Tx group than in the NTx group (Figure 2J-K; \**p*<0.05). Interestingly, SRB1 was expressed only in the theca cell layer of follicles within the ovarian tissue (Figure 2L). The level of SRB1 in ovarian follicles was significantly lower in the NTx group than in the normal group and significantly greater in the Tx group than in the NTx group (Figure 2M; \**p*<0.05).

### PD-MSC transplantation decreases vascular permeability in the ovaries of TAA-injured rats

SRB1 is known to activate endothelial nitric oxide synthase (eNOS), and the ornithine pathway is integral to endothelial cell metabolism and works in conjunction with eNOS to regulate vascular homeostasis ^37^. In the ornithine pathway, eNOS facilitates the production of nitric oxide and citrulline, both of which are critical for endothelial function, promoting vasodilation and maintaining vascular homeostasis by regulating blood flow and vessel tone (Figure 3A). The level of citrulline in individual serum samples from TAA- injured rats was significantly lower in the NTx group than in the normal group. However, compared with that in the NTx group, the citrulline level in individual serum samples from TAA-injured rats substantially increased after PD-MSC transplantation (Figure 3B; \**p*<0.05). The ratio of eNOS dimers to monomers was lower in the NTx group than in the normal group but was greater in the Tx group than in the NTx group (Figure 3C). These results suggest that citrulline is produced through eNOS coupling, thereby regulating vascular homeostasis.

**FIGURE 3.**
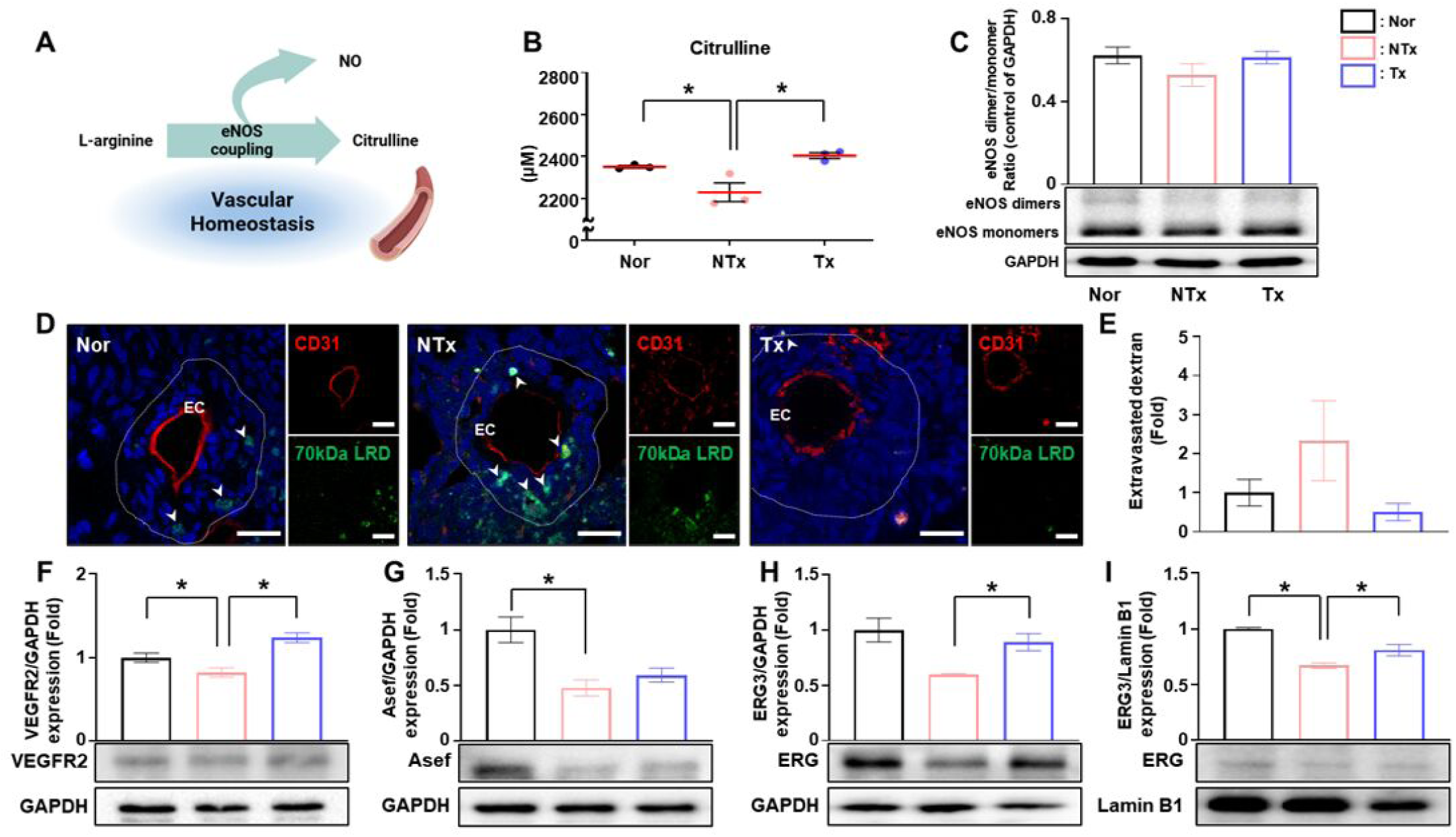
Effect of PD-MSCs on vascular permeability in ovaries with lipotoxicity. (a) Schematic of the mechanism by which the ornithine cycle contributes to vascular homeostasis. (b) Secretion of citrulline in the serum was analyzed via ELISAs. (c) The protein expression of eNOS was analyzed by low-temperature SDS‒PAGE. (d) Leakage of 70 kDa rhodamine dextran was analyzed in the blood vessels of the ovarian tissues. Magnification: 63X; scale bar: 20 μm. (e) The intensity of the 70 kDa rhodamine dextran-positive signal versus the CD31-positive signal was quantified via the ImageJ program. (f) The protein expression of VEGFR2, (g) Asef and (h) ERG in total lysates of ovarian tissues was analyzed by western blotting. (i) The protein expression of ERG in the nuclear fraction of ovarian tissues was analyzed by western blotting. * *p*<0.05. The *p* value was determined via a nonparametric Kruskal‒Wallis test followed by Conover‒Iman’s multiple comparisons with BH correlation.

For analysis of vascular permeability in the microvasculature of the ovaries of TAA-injured rats, rhodamine 70 kDa, which can be used to analyze blood flow leakage, was administered before the rats were sacrificed (Figure 3D). The intensity of extravasated dextran in the ovarian microvasculature was substantially greater in the NTx group than in the normal group and significantly lower in the Tx group than in the NTx group (Figure 3E; \**p*<0.05).

To determine vascular permeability in the ovaries of TAA-injured rats, we analyzed factors related to vascular permeability. The protein expression of VEGFR2, which is known as the vascular permeability factor (VPF; VEGF) receptor, was significantly lower in the NTx group than in the normal group but considerably greater in the Tx group than in the NTx group (Figure 3F; \**p*<0.05). The protein expression of Rho guanine nucleotide exchange factor 4 (Asef), which is known to regulate endothelial cell permeability, was significantly lower in the NTx group than in the normal group but was greater in the Tx group than in the NTx group (Figure 3G; \**p*<0.05). The protein expression of ETS transcription factor (ERG) in the total lysate of ovarian tissues was lower in the NTx group than in the normal group but significantly greater in the Tx group than in the NTx group (Figure 3H; \**p*<0.05). The protein expression of ERG in the nuclear fraction of ovarian tissues was significantly lower in the NTx group than in the normal group but significantly greater in the Tx group than in the NTx group (Figure 3I; \**p*<0.05). These results indicate that vascular permeability is reduced in the ovarian tissues of TAA-injured rats following PD-MSC transplantation, likely through the activation of eNOS.

### PD-MSC transplantation promotes steroidogenesis in the ovaries of TAA-injured rats

ApoA1 binds to mature HDL via ATP binding cassette subfamily A member 1 (ABCA1) and then interacts with the SRB1 receptor to carry out steroidogenesis (Figure 4A). The secretion level of ABCA1, which delivers lipids to ApoA1 to form mature HDL, was significantly lower in the NTx group than in the normal group but significantly greater in the Tx group than in the NTx group in individual serum samples (Figure 4B; \**p*<0.05). The protein expression of ABCA1 in ovarian tissues was substantially lower in the NTx group than in the normal group and significantly greater in the Tx group than in the NTx group (Figure 4C; \**p*<0.05). In addition, the localization and expression of ABCA1 in mature follicles of ovarian tissues were dramatically lower in the NTx group than in the normal group. Compared with that in the NTx group, this factor was substantially greater in the Tx group (Figure 4D-E; \**p*<0.05).

**FIGURE 4.**
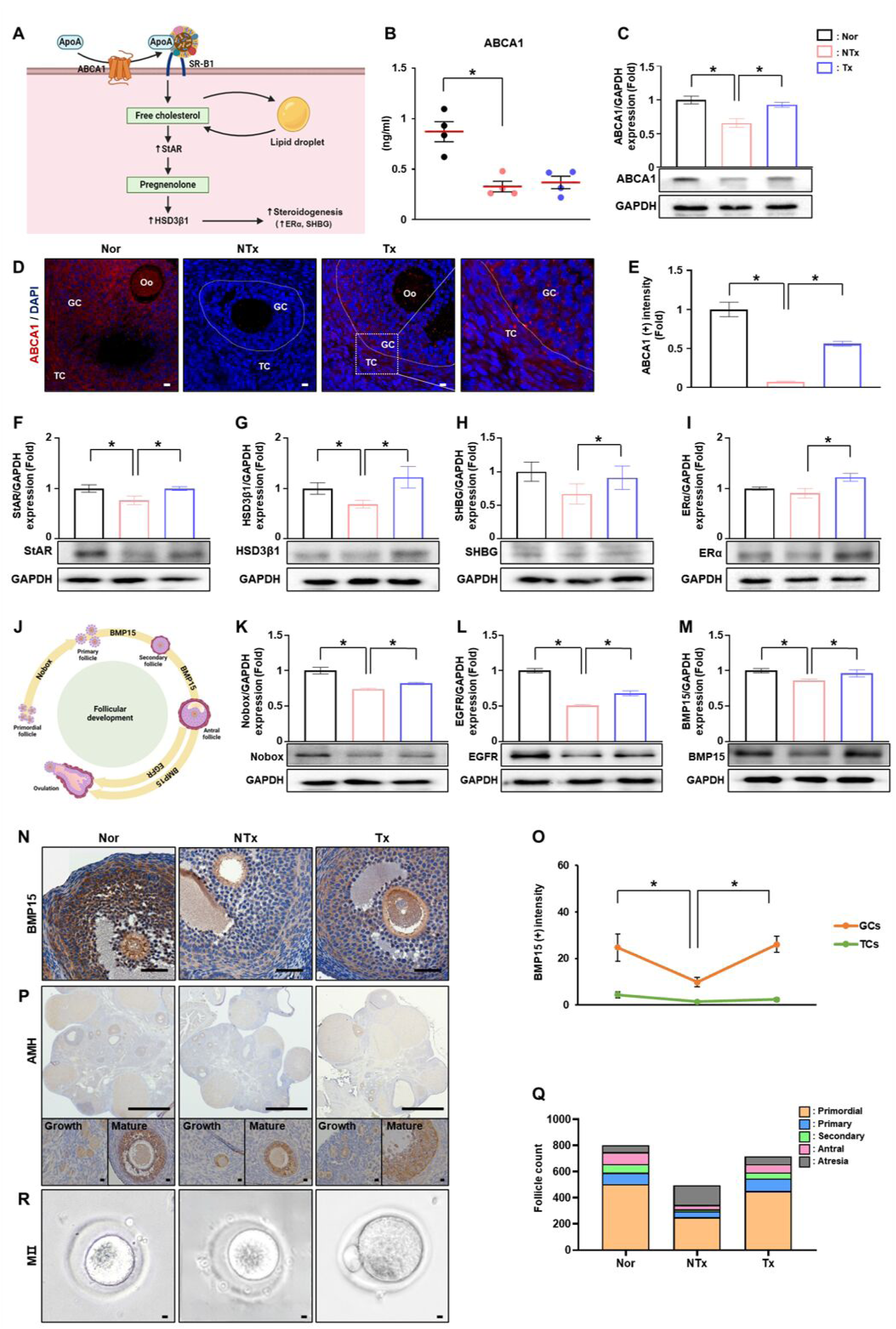
Effect of PD-MSCs on follicular development by steroidogenesis in ovaries with lipotoxicity. (a) Mechanism of ApoA1-SRB1 signaling in steroidogenesis. (b) ABCA1 secretion was analyzed in the serum of TAA-treated rats. (c) ABCA1 protein expression was analyzed by western blotting. (d) ABCA1 localization was visualized by immunofluorescence staining. (e) The intensity was quantified via the ImageJ program. Magnification: 20X; scale bar: 50 μm. (f) The protein expression of StAR, (g) HSD3β1, (h) SHBG and (i) ERα was analyzed by western blotting. (j) Schematic of genes involved in each stage of follicular development. (k) The protein expression of Nobox, (l) EGFR and (m) BMP15 was analyzed by western blotting. (n) The localization of BMP15 in ovarian tissues was visualized by immunohistochemical staining. (o) The intensity of the BMP15-positive signal in granulosa cells and theca cells was quantified via the 3D HISTECH program. Magnification: 40X; scale bar: 50 μm. (p) The localization of AMH was visualized by immunohistochemical staining. Magnification: 1X and 40X; scale bar: 50 and 500 μm. (q) Follicles from each stage were analyzed via the 3D HISTECH program. (r) Oocytes from all groups were collected after superovulation. * *p*<0.05. The *p* value was determined via a nonparametric Kruskal‒Wallis test followed by Conover‒Iman’s multiple comparisons with BH correlation.

To determine whether steroidogenesis is activated, we analyzed the expression of steroidogenesis-related factors in the ovaries of TAA-injured rats. Compared with that in normal ovarian tissues, the protein expression of StAR, which delivers cholesterol to the inner mitochondrial membrane and regulates steroid hormone biosynthesis, was significantly lower in the NTx group. Compared with that in the NTx group, the expression of this gene in ovarian tissues was significantly greater in the Tx group (Figure 4F; \**p*<0.05). The protein expression of HSD3β1, which transports cholesterol into mitochondria via StAR, was lower in the NTx group than in the normal group but was greater in the Tx group than in the NTx group in ovarian tissues (Figure 4G; \**p*<0.05). The protein expression of sex hormone binding globulin (SHBG) and estrogen receptor alpha (ERα) was lower in the NTx group than in the normal group but significantly greater in the Tx group than in the NTx group (Figure 4H-I; \**p*<0.05). These results indicate that PD-MSCs promote steroidogenesis in ovaries with metabolic disorders.

### PD-MSC transplantation protects follicular development in the ovaries of TAA-injured rats

To demonstrate that follicular development was protected by decreased vascular permeability, we examined the protein expression of genes related to follicular development in the ovarian tissues of TAA-injured rats (Figure 4J). The protein expression of Nobox, which is essential for early follicle development, was significantly lower in the NTx group than in the normal group but significantly greater in the Tx group than in the NTx group in ovarian tissues (Figure 4K; \**p*<0.05). The protein expression of epidermal growth factor receptor (EGFR) and bone morphogenetic protein 15 (BMP15), which are related to follicular maturation, was considerably lower in the NTx group than in the normal group, and their expression was significantly greater in the Tx group than in the NTx group (Figure 4I-M; \**p*<0.05). In particular, BMP15 was localized in mature follicles (Figure 4N), and the intensity of BMP15 in the granulosa cells of mature follicles was markedly lower in the NTx group than in the normal group. The intensity of this molecule in the granulosa cells of mature follicles was substantially greater in the Tx group than in the NTx group. However, BMP15 expression in theca cells of mature follicles did not differ among the groups (Figure 4O; \**p*<0.05).

For analysis of follicular development at different stages, AMH, which is expressed by granulosa cells of developing follicles, was detected in the ovaries of TAA-injured rats. In the NTx group, fewer primordial follicles and smaller mature follicles (e.g., secondary, antral follicles) were observed (Figure 4P). Additionally, to assess follicular development at different stages, we performed follicle counting on more than three people after H&E staining. In the NTx group, the number of follicles at each stage (e.g., primordial, primary, secondary and antral follicles) was significantly lower than that in the normal group. Specifically, mature follicles (e.g., secondary and antral follicles) did not progress in the NTx group but matured when PD-MSCs were transplanted. Additionally, we found a notable significant increase in atretic follicle number in the NTx group compared with the normal group and a significant decrease in the Tx group compared with the NTx group (Figure 4Q, Table 1; \**p*<0.05). These results indicate that PD-MSC transplantation protected follicular development and supported granulosa cell maturation in ovaries with metabolic disorders.

### PD-MSC cocultivation decreases vascular permeability in TAA-treated human umbilical vein endothelial cells (HUVECs) (*in vitro*)

To demonstrate lipid accumulation in endothelial cells, we conducted an *in vitro* experiment by treating HUVECs with TAA, a substance that induces lipid toxicity, and stained them with BODIPY 505/515 (Figure 5A). Compared with those in the control group, the number and area of lipid droplets significantly increased in the TAA-treated group but significantly decreased in the TAA-treated group cocultivated with PD-MSCs compared with those in the TAA-treated group (Figure 5B-C; \**p*<0.05).

**FIGURE 5.**
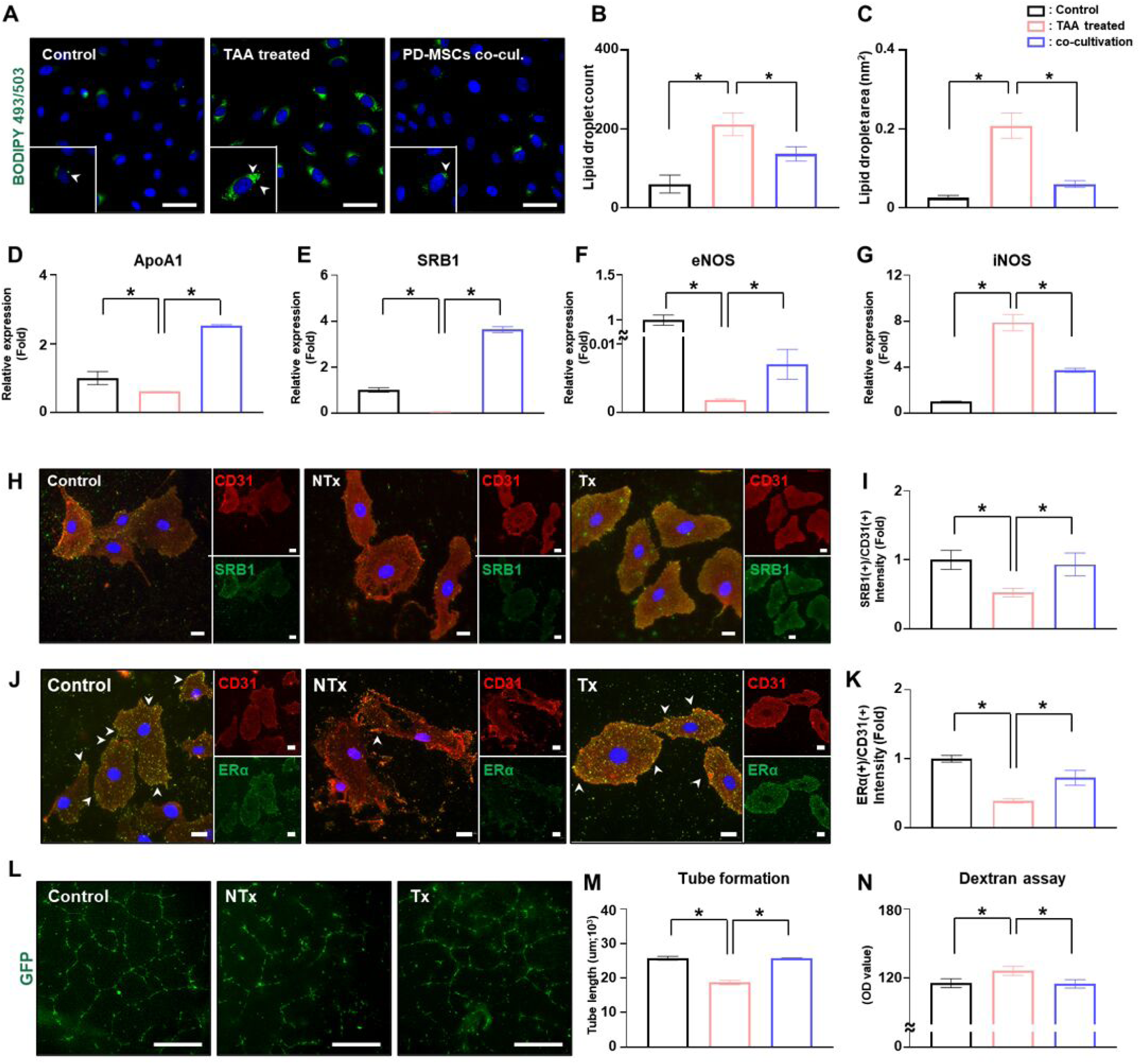
Effect of PD-MSCs on vascular permeability by ApoA1-SRB1 signaling in TAA-treated HUVECs *in vitro*. (a) Lipid droplets in TAA-treated HUVECs were visualized by BODIPY 505/515 staining. (b-c) The count and area of lipid droplets were quantified via the ImageJ program. Magnification: 40X; scale bar: 50 μm. (d) The mRNA expression of ApoA1, (e) SRB1, (f) eNOS and (g) iNOS was analyzed by qRT‒PCR. (h) The localization of SRB1 was visualized by immunofluorescence staining, and endothelial cells were labeled with CD31. (i) The intensity of SRB1 versus CD31 was quantified via the ImageJ program. (j) The localization of ERα was visualized by immunofluorescence staining. (k) The intensities of ERα and CD31 were quantified via the ImageJ program. Magnification: 40X; scale bar: 50 μm. (l) Tube formation of HUVECs was visualized by DiI staining. (m) Total length of tube formation was quantified via the ImageJ program. Magnification: 4X; scale bar: 500 μm. (n) Vascular permeability was analyzed by a dextran assay. * *p*<0.05. The *p* value was determined via a nonparametric Kruskal‒Wallis test followed by Conover‒Iman’s multiple comparisons with BH correlation.

We investigated whether this effect was strengthened through the activation of ApoA-SRB1 signaling by PD-MSCs. Compared with that in the TAA-treated group, the ApoA1 mRNA expression in the TAA-treated group cocultivated with PD-MSCs was significantly greater (Figure 5D). The mRNA expression of SRB1 and eNOS was substantially lower in the TAA-treated group than in the normal group but notably increased in the PD-MSC cocultivation with TAA treatment group (Figure 5E-F; \**p*<0.05). However, the mRNA expression of inducible nitric oxide synthase (iNOS), which promotes vascular permeability by excessively producing NO, was markedly increased in the TAA-treated group compared with the normal group but significantly decreased in the PD-MSC cocultivation with TAA treatment group (Figure 5G; \**p*<0.05). In particular, the expression of SRB1 was significantly lower in the TAA-treated group than in the normal group but was significantly greater in the PD-MSC cocultivation with TAA treatment group (Figure 5H-I; \**p*<0.05).

To verify vascular permeability in endothelial cells by ApoA-SRB1 signaling, we treated HUVECs with TAA. Low estrogen levels increase vascular permeability, which is closely associated with atherosclerosis ^38^. Compared with those in the control HUVECs, the localization and expression of ERα in the HUVECs treated with TAA were notably reduced. However, the expression was significantly greater in the PD-MSC cocultivation with TAA treatment group than in the TAA-treated group (Figure 5J-K; \**p*<0.05).

The capacity for endothelial cell tube formation was impaired, with bridges and branch points disrupted in several areas when TAA was added. However, when PD-MSCs were cocultivated, the bridges reformed, and the tube length significantly increased (Figure 5L‒M; \**p*<0.05). Furthermore, we assessed vascular permeability using dextran. Vascular permeability was significantly greater in the TAA-treated group than in the control group, but cocultivation with PD-MSCs significantly decreased vascular permeability compared with that in the TAA-treated group (Figure 5N).

### PD-MSC cocultivation increases steroidogenesis in SRB1 inhibitor- or TAA-treated primary theca cells (*in vitro*)

To investigate whether ApoA1-SRB1 signaling through PD-MSCs affects ovarian follicles and to understand the underlying mechanism, we treated theca cells, which specifically express SRB1, with the SRB1 inhibitor BLT-1, as shown in Figure 2I. The ratio of ApoB to ApoA1 in the supernatant of primary theca cells was significantly greater in the BLT-1-treated group than in the control group but was significantly lower in the PD-MSC cocultivation with BLT-1 treatment group (Figure 6A; \**p*<0.05). Compared with that in the control group, the level of ABCA1 in the supernatant of primary theca cells was markedly lower in the BLT-1-treated group, but cocultivation with PD-MSCs increased the ABCA1 level compared with that in the BLT-1-treated group (Figure 6B; \**p*<0.05). Interestingly, the levels in the TAA-treated groups were similar to those in the BLT-1-treated groups. These results indicate that the expression of SRB1 is suppressed even after treatment with TAA. Next, we analyzed whether PD-MSCs activate SRB1. Compared with that in the control group, the expression of SRB1 in primary theca cells was lower in both the BLT-1-treated and TAA-treated groups, but an increase was observed in the PD-MSC cocultivation with BLT-1 or TAA groups compared with the respective treatment groups (Figure 6C-D; \**p*<0.05). Compared with that in the control group, the protein expression of SRB1 was decreased in the BLT-1-treated group but notably increased in the PD-MSC cocultivation with BLT-1 treatment group compared with the BLT-1-treated group (Figure 6E; \**p*<0.05). These results indicate that PD-MSCs activate ApoA-SRB1 signaling.

**FIGURE 6.**
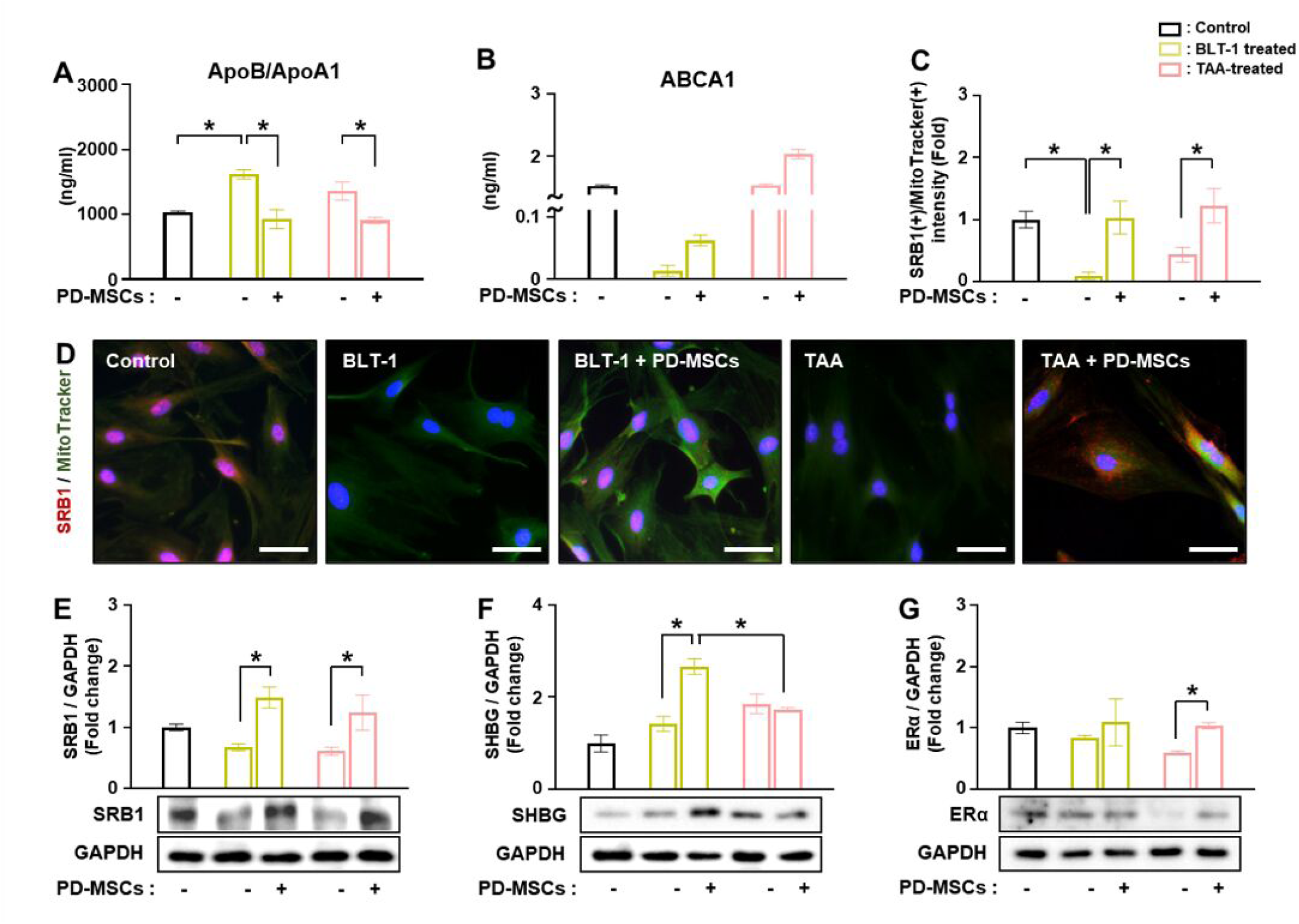
Effect of PD-MSCs on steroidogenesis in TAA- or BLT-1-treated primary theca cells *in vitro*. (a) The ratio of ApoB to ApoA1 and (b) secretion of ABCA1 were analyzed in the supernatants of primary theca cells via ELISAs. (c-d) The localization of SRB1 was visualized by immunofluorescence staining, and the intensity of SRB1 was quantified via the ImageJ program. Magnification: 40X; scale bar: 50 μm. (e) The protein expression of SRB1, (f) SHBG and (g) ERα was analyzed by western blotting. * *p*<0.05. The *p* value was determined via a nonparametric Kruskal‒Wallis test followed by Conover‒Iman’s multiple comparisons with BH correlation.

To determine whether ApoA1-SRB1 signaling is involved in steroidogenesis, we analyzed steroidogenesis-related factors in BLT-1-treated primary theca cells. SHBG protein expression was significantly greater in the PD-MSC cocultivation with BLT-1 treatment group than in the BLT-1-treated group (Figure 6F; \**p*<0.05). Compared with that in the control group, the protein expression of ERα was decreased in both the BLT-1-treated and TAA-treated groups but was increased in the cocultivation groups compared with the respective treatment groups (Figure 6G; \**p*<0.05). These results show that PD-MSCs increased the steroidogenesis by ApoA-SRB1 signaling in primary theca cells.

## DISCUSSION

Dysregulation of lipid metabolism leads to excessive lipid accumulation, triggering lipotoxicity, which induces oxidative stress, endoplasmic reticulum (ER) stress, and mitochondrial dysfunction in cells. As key regulators of ATP production and metabolic control, mitochondria play crucial roles in the pathophysiology of PCOS by maintaining energy homeostasis in ovarian cells ^39^. In PCOS, mitochondrial dysfunction leads to reduced ATP synthesis, impairing steroidogenesis. This disruption in mitochondrial function exacerbates insulin resistance, hormonal imbalances, and follicular dysfunction, which are hallmark features of PCOS ^40^. Recent reports indicate that mitochondria play a key role in the pathogenesis of metabolic diseases such as obesity and type 2 diabetes ^41^. The damage to and degradation of mitochondrial components strongly contribute to the aging process.

Additionally, atherosclerosis is closely linked to aging, as this condition becomes more widespread and severe with advancing age ^42^. Unlike previous studies that focused solely on lipid accumulation as a consequence of metabolic disorders, our study suggests that regulating lipid accumulation within blood vessels could directly influence vascular function and contribute to delaying aging. These reports suggest that alleviating lipid accumulation within blood vessels may not only improve functionality but also contribute to rejuvenation during the aging process.

In women with PCOS, there are various vascular abnormalities in the ovaries (e.g., arterial hyperemia and vascular dysfunction). These changes occur due to an imbalance in vascular angiogenic factors that regulate vascular function, leading to abnormal follicular development through the formation of abnormal blood vessels ^13^. Takehara and colleagues demonstrated that the CD34-positive signal increases in the ovaries of cyclophosphamide-induced rats transplanted with AD-MSCs, since AD-MSCs secrete vascular angiogenic cytokines (e.g., VEGF, IGF, and HGF) ^43^. Xia and colleagues reported that BM-MSCs reduce the degree of primordial follicle apoptosis by increasing the microvascular density in ovaries^44^. However, previous studies predominantly emphasized vascular angiogenesis without considering vascular homeostasis, which is crucial for ovarian function. The limitation of most studies addressing improvements in abnormal ovarian vasculature is their focus solely on vascular angiogenesis. Vascular angiogenesis within the ovaries is closely associated with ovarian cancer ^45^. In our previous report, we demonstrated that while the number of blood vessels in the ovaries of rats transplanted with PD-MSCs did not differ, vascular permeability decreased because of the activation of cytokines such as PDGF and HGF ^26, 46^. For vascular homeostasis, a balance of the following factors is necessary: (1) vasodilation, (2) actin formation, (3) vascular permeability, (4) oxidative stress, and (5) inflammation rather than angiogenesis alone ^47^. This study represents the first report demonstrating that ovarian vascular homeostasis is regulated by the activated ApoA1-SRB1-eNOS pathway induced by PD-MSCs.

Endothelial cell metabolism disorders have been shown to affect several pathways of central carbon metabolism ^48^. Vascular homeostasis is regulated by the production of nitric oxide (NO) through the ornithine pathway in endothelial cell metabolism pathways ^49, 50^. Under normal conditions, eNOS converts L-arginine into citrulline and NO. In the absence of NO production, eNOS can become uncoupled and generate superoxide anions (O2•^−^), which can react with NO to form peroxynitrite (ONOO^−^), a highly reactive oxygen species. eNOS uncoupling leads to the generation of superoxide instead of NO, which serves as a source of harmful free radicals ^51^. The uncoupling of eNOS is causally linked to metabolic diseases. Milewski and colleagues demonstrated that the eNOS dimer/monomer ratio significantly decreased in the brain cortex of TAA-treated rats with acute liver failure ^52^. In our study, we observed a significant reduction in the eNOS dimer/monomer ratio in the ovaries of the TAA-treated group, whereas PD-MSC transplantation restored this ratio to a level comparable to that of the normal group, further confirming the role of PD-MSCs in vascular protection. Moreover, HDL binding to SR-B1 induces cholesterol efflux and activates eNOS ^53^. NO synthesis is associated not only with vascular homeostasis but also with ovarian function in individuals with PCOS. NO has also been implicated in the regulation of gonadotropin secretion at the level of the ovarian follicle barrier, where it is found at the level of the ovarian microvasculature ^54^. In women with PCOS, the reduced production of NO due to decreased iNOS/eNOS leads to endothelial cell dysfunction ^55^. Interestingly, eNOS is detected in the theca cell layer, ovarian stroma, and surface of oocytes, where NO induces ovulation and reduces the apoptosis of follicular cells in normal ovaries ^13^. To further evaluate the impact of PD-MSC transplantation on ovulatory dysfunction caused by TAA-induced lipid metabolism abnormalities, we performed superovulation and collected oocytes to assess their quality and functional potential (Figure 4R).

Clinical studies as well as preclinical studies on PCOS are currently underway. According to the NIH Clinical Trials Database (www.clinicaltrials.gov), numerous clinical trials investigating PCOS are currently in progress. Most clinical trials for PCOS are focused on metformin, a drug that regulates insulin sensitivity. Pharmacotherapy for PCOS (e.g., metformin, combined oral contraceptive pills, and clomiphene citrate) alleviates insulin sensitivity or hormonal imbalances, but these effects are only temporary. Additionally, side effects such as digestive issues and an increased risk of cardiovascular disease have been reported ^56^. Therefore, treatments with a precise mode of action (MoA) for PCOS, such as regulating cholesterol lipid metabolism and normalizing vascular endothelial function, are needed. MSCs have been shown to promote therapeutic efficacy in degenerative diseases through their paracrine effect, with several studies reporting this MoA. MSC transplantation exerts its paracrine effects by reaching damaged tissues and restoring the microenvironment through immunomodulation, tissue regeneration and healing, antifibrosis, antiapoptosis, and angiogenesis, thereby promoting tissue repair ^57^. However, clinical trials involving adult stem cell therapy are currently relatively limited. While some clinical trials have identified alleviation of insulin resistance following UC-MSC transplantation, these studies remain in early phases, emphasizing the need for larger-scale trials to validate the efficacy and safety of MSCs.

In conclusion, our findings indicated that PD-MSCs activated ApoA-SRB1 signaling. After PD-MSC transplantation into rats with metabolic disorders, lipid accumulation and vascular permeability decreased. Moreover, PD-MSC transplantation normalized ovarian function by increasing steroidogenesis in the ovaries of rats with metabolic disorders. These findings provide strong evidence that PD-MSCs contribute to ovarian vascular function by regulating endothelial function and lipid metabolism. Therefore, these findings offer new insights into stem cell therapy for reproductive medicine and highlight the potential of PD- MSCs as a novel therapeutic approach for PCOS-related infertility.

## Data Availability

Yes, it could be.

## ACKNOWLEDGEMENTS

The authors would like to thank Ms. Young Shin Lee (University of CHA, Gyeonggi-do, Korea) for the graphical abstract, and the illustration was created with https://app.biorender.com/. Additionally, we sincerely thank Mr. Jun Hyeong You, M.S., for his valuable assistance in this research.

## FOOTNOTE

### Nonstandard Abbreviations and Acronyms

PD-MSCs: placenta-derived mesenchymal stem cells
TAA: thioacetamide
Tx: transplantation
NTx: non-transplantation
ApoA1: apolipoprotein A1
SRB1: scavenger receptor class B member 1
HOMA-IR: homeostatic model assessment for insulin resistance
PCOS: polycystic ovary syndrome
HDL-C: high-density lipoprotein cholesterol
LDL-C: low-density lipoprotein cholesterol
eNOS: endothelial nitric oxide synthase
AMH: anti-Mullerian hormone
E2: estradiol
LH: luteinizing hormone
FSH: follicle stimulating hormone
TES: testosterone

## ETHICS APPROVAL AND CONSENT TO PARTICIPATE

All animal experiments were approved by the Institutional Animal Care and Use committee (IACUC 220044) of the CHA Laboratory Animal Research Center at Sampyeong-dong in Gyeonggi, Republic of Korea. Also, the collection of human placenta and their use were conducted under the guidelines and with the approval of the affiliated Institutional Review Board of the Gangnam CHA General Hospital, Seoul, Korea (IRB 07-18). All patients provided written informed consent to the respective use of their tissues.

## AUTHOR CONTRIBUTIONS

For research articles with several authors a short paragraph specifying their individual contributions must be provided. The following statements should be used “Conceptualization, G.J.K.; Animal model construction, validation and formal analysis, H.P., D.H.L., J.H.Y., H.W.J., J.Y.S., J.U.N.; Cell culture, H.P.; Image data analysis, H.P., J.H.Y.; Writing and editing, H.P., J.S., H.L. and G.J.K.; project administration, and funding acquisition, G.J.K.” All authors have read and agreed to the published version of the manuscript.

## DATA AVAILABILITY STATEMENT

The data that support the findings of this study are available from the corresponding author upon reasonable request.

## SUPPORTING INFORMATION

Additional supporting information may be found online in the supporting information section.

